# Liver Chemistries in Patients with Severe or Non-severe COVID-19: A Meta-Analysis

**DOI:** 10.1101/2020.04.24.20074179

**Authors:** Xuan Dong, Dan-Yi Zeng, Yan-Yan Cai, Wei-Ming Chen, Qing-Qing Xing, Yan-Dan Ren, Mei-Zhu Hong, Jin-Shui Pan

**Author notes:** E-mails: Xuan Dong. These authors contributed equally to this work (Xuan Dong, Dan-Yi Zeng, Yan-Yan Cai). Correspondence: Jin-Shui Pan, MD, PhD, Liver Research Center, the First Affiliated Hospital of Fujian Medical University, No. 20 Chazhong Road, Fuzhou 350001, Fujian, China., Mei-Zhu Hong, MD, Department of Traditional Chinese Medicine, Zhongshan Hospital Affiliated to Xiamen University, 201 Hu-Bin Nan Road, Xiamen, Fujian 361004, China.

## Abstract

**Background and Aims:** Cumulating observations have indicated that patients with coronavirus disease (COVID-19) undergo different patterns of liver impairment. We performed a meta-analysis of published liver manifestations and described the liver damage in COVID-19.

**Methods:** We searched PubMed, Google Scholar, Embase, Cochrane Library, medRxiv, bioRxiv, and three Chinese electronic databases through April 18, 2020, in accordance with the Preferred Reporting Items for Meta-Analyses. We analyzed pooled data on liver chemistries stratified by COVID-19 severity using a fixed or random-effects model.

**Results:** In the meta-analysis of 37 studies, which included a total of 6,235 patients, the pooled mean alanine aminotransferase (ALT) was 36.4 IU/L in the severe COVID-19 cases and 27.8 IU/L in the non-severe cases (95% confidence interval [CI]: − 9.4 to − 5.1, p < 0.0001). The pooled mean aspartate aminotransferase (AST) was 46.8 IU/L in the severe cases and 30.4 IU/L in the non-severe cases (95% CI: − 15.1 to − 10.4, p < 0.0001). Furthermore, regardless of disease severity, the AST level is often higher than the ALT level. Compared with the non-severe cases, the severe cases tended to have higher γ-glutamyltransferase levels but lower albumin levels.

**Conclusions:** In this meta-analysis, we comprehensively described three patterns of liver impairment related to COVID-19, namely hepatocellular injury, cholestasis, and hepatocellular disfunction, according to COVID-19 severity. Patients with abnormal liver test results are at higher risk of progression to severe disease. Close monitoring of liver chemistries provides an early warning against disease progression.

**Lay Summary:** Data on abnormal liver chemistries related to coronavirus disease (COVID-19) are cumulating but are potentially confusing. We performed a meta-analysis of 37 studies that included a total of 6,235 patients with COVID-19. We noted that patients with abnormal liver test results are at higher risk of progression to severe disease and close monitoring of liver chemistries provides early warning against disease progression.

## INTRODUCTION

According to the situation report released by the World Health Organization (WHO), as of April 18, 2020, 2,160,207 coronavirus disease (COVID-19) cases were confirmed globally, of which 146,088 led to deaths [1]. Although effectively controlled in mainland China, the spread and upward trend of COVID-19 have accelerated dramatically worldwide. In response to the emerging threat posed by COVID-19, caused by severe acute respiratory syndrome coronavirus 2 (SARS-CoV-2), the WHO has declared a Public Health Emergency of International Concern on January 30, 2020, which was further labeled as a pandemic on March 11, 2020. Similarly, the other two previously identified coronaviruses, namely SARS-CoV and Middle East Respiratory Syndrome-CoV, SARS-CoV-2 causes severe viral pneumonia in humans. As no specific acquired immunity exists in the general population, SARS-CoV-2 has high infectivity, which has resulted in an ongoing global health crisis.

Apart from the respiratory system, the urinary system, gastrointestinal tract, and even the central nervous system are the probable target organs of SARS-CoV-2, which utilizes the angiotensin-converting enzyme 2 (ACE2) receptors of the respiratory and gastrointestinal tracts as the entry point for epithelial cells [2]. Among patients’ common complaints related to COVID-19 are gastrointestinal symptoms, including nausea/vomiting, diarrhea, and abdominal pain [3-6]. Abundant ACE2 protein expression in the glandular cells of gastric, duodenal, and rectal epithelia supports the entry of SARS-CoV-2 into the host epithelial cells [7]. Single-cell RNA sequencing has revealed a specific ACE2 expression in cholangiocytes [8]. Thus, performing liver chemistry tests for a number of patients with COVID-19 seems reasonable. In fact, several studies have found liver injury in patients with COVID-19 [9-13].

Furthermore, cumulating observations have indicated that different patterns of abnormal liver chemistries exist between patients with severe and non-severe COVID-19. Although liver manifestations of COVID-19 pose an important diagnostic challenge to clinicians when treating patients with symptoms related to COVID-19 on initial presentation, these are potentially useful for recognizing severe cases of COVID-19 in the early stage.

Considering the diverse clinical manifestations and increasing number of reported COVID-19 cases, a systematic summary of the liver manifestations of COVID-19 is urgently needed.

Liver chemistries generally consist of hepatocellular injury-related indexes, including alanine aminotransferase (ALT) and aspartate aminotransferase (AST); cholestatic injury-related indexes, comprised of alkaline phosphatase (AKP) and γ-glutamyltransferase (GGT); and hepatocellular function-related indexes such as albumin (ALB) level and prothrombin time (PT)^[14]^. In general, total bilirubin (TBIL), direct bilirubin (DBIL), and globulin (GLB) levels, and international standardized ratio (INR) are also assessed in clinical practice. However, observations to comprehensively analyze liver chemistries in patients with COVID-19 patients are rather few. Hence, the aim of this study was to provide a comprehensive view of liver test parameters in patients with severe and non-severe COVID-19. With better knowledge, more targeted therapies and holistic care approaches could be developed.

## METHODS

### Studies selection

The following databases were searched from December 1, 2019, through April 18, 2020: PubMed, Google Scholar, Embase, Cochrane Library, medRxiv, bioRxiv, and three Chinese electronic databases (CQVIP, Wanfang Data, and Chinese National Knowledge Infrastructure). “Coronavirus,” “COVID-19,” “2019-nCoV-2,” “SARS-CoV-2,” or novel coronavirus were used as search keywords. Potential studies were retrieved in accordance with the PRISMA guideline [15]. Details of the database search are listed in the Supplementary file. The retrieved articles were imported to Endnote X9.3 (Thompson and Reuters, Philadelphia, Pennsylvania), and duplicates were removed.

### Selection criteria

The eligibility of the potential studies was determined independently by two authors (XD and WMC), and dissonance was arbitrated by the third author (JSP). The inclusion criteria were as follows: (1) study population: adult COVID-19 patients; (2) study design: case series, case report, prospective cohort study, retrospective cohort study, case-control study, and randomized controlled trial; and (3) language: studies published in English or Chinese. The exclusion criteria were as follows: (1) pediatric patients or pregnant women; (2) patients without nucleic acid data or serology evidence of SARS-CoV2 infection; (3) asymptomatic patients with SARS-CoV2 infection; (3) study design: review article, meta-analysis, editorial, or commentary. Studies that only reported the percentages of the indexes related to liver chemistries rather than the mean or median values of the corresponding indexes were also excluded.

### Data extraction

For the eligible articles, we recorded the following items: first author, study location, sample size, patient age and sex, and liver chemistry-related indexes such as ALT, AST, TBIL, DBIL, GGT, AKP, and ALB levels. The severity of COVID-19 was also recorded.

### Data analysis

The statistical analyses were performed using the R version 3.2.3 statistical software (R Foundation for Statistical Computing). The continuous variables that showed a normal distribution were expressed as mean ± SD, while those that conformed to a skewed distribution were expressed as median (interquartile range [IQR]). For the studies that provided summary data of median, minimum, and maximum values, we used the method developed by Luo et al [16] to estimate the sample mean and SD for the continuous outcomes. The online tool used is provided at http://www.math.hkbu.edu.hk/~tongt/papers/median2mean.html. The 95% confidence interval (CI) was presented as a Forest plot. The Cochran Q test was used to detect the heterogeneity among studies, with a p value of <0.10 indicating significant heterogeneity. The *I*^2^ statistics was calculated to measure the proportion of total variation among the studies to which the heterogeneity was attributed. *I*^2^ values of <25%, 25–75%, and >75% represent low, moderate, and high heterogeneity, respectively [17]. Publication bias was evaluated using a funnel plot. A subgroup analysis was performed according to disease severity.

## RESULTS

### Characteristics of the studies included in the meta-analysis

The selection process of the potential studies is depicted in Figure 1. Of the 2,127 studies identified, 37 studies were included in the meta-analysis. The characteristics of the enrolled studies are listed in Table 1 [9, 12, 13, 18-51]. Information, including the study location, sample size, patient age and sex, disease severity, INR, PT, and TBIL, DBIL, ALT, AST, GGT, AKP, ALB, and GLB levels, was recorded. The mean ages of patients with non-severe and severe COVID-19 were 51.1 and 64.0 years, respectively (Supplementary Figure 1). In the enrolled studies, male patients accounted for 54.6%. Among the studies that reported disease severity, severe disease accounted for 30.3% (range, 8.0–54.8%) of the cases.

**Table 1.**
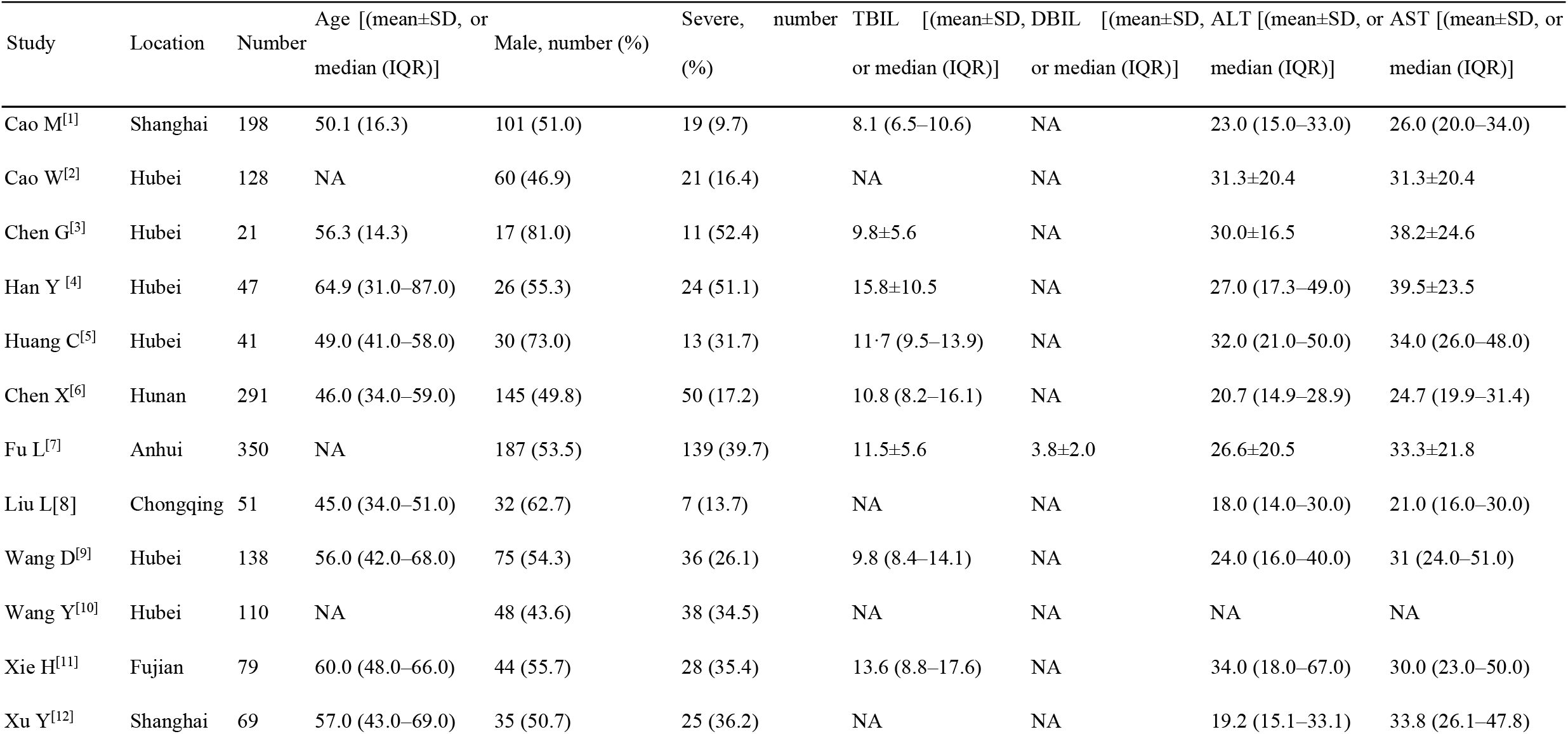
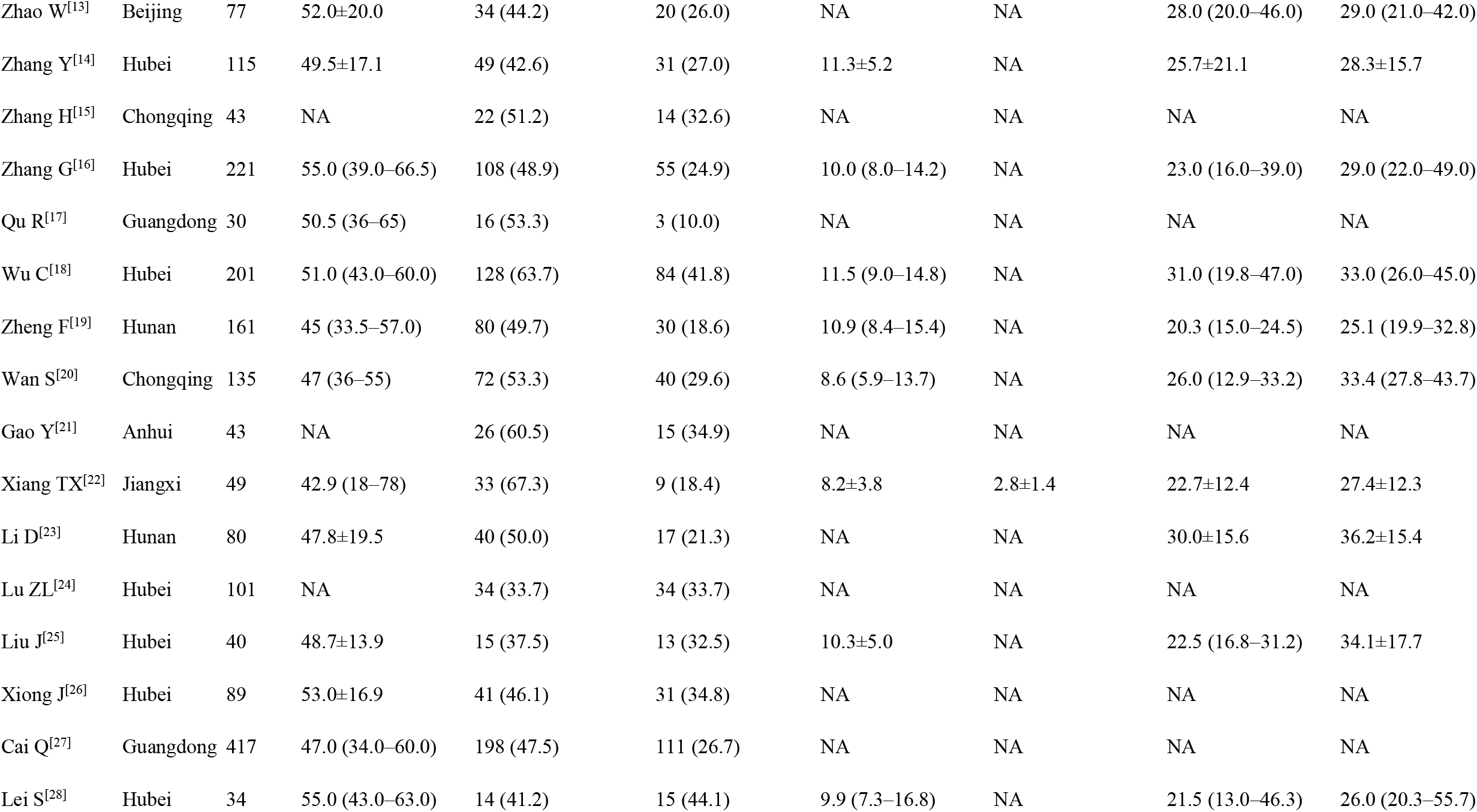
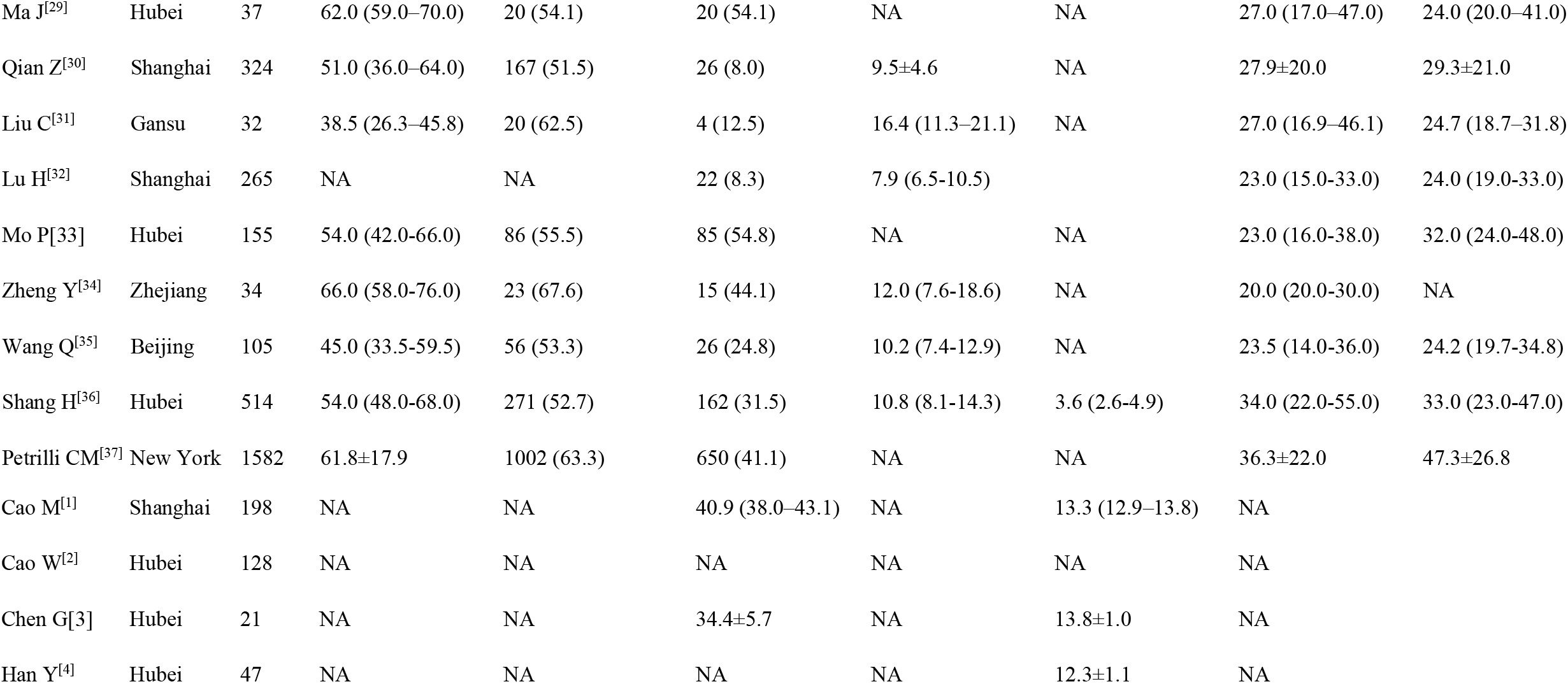
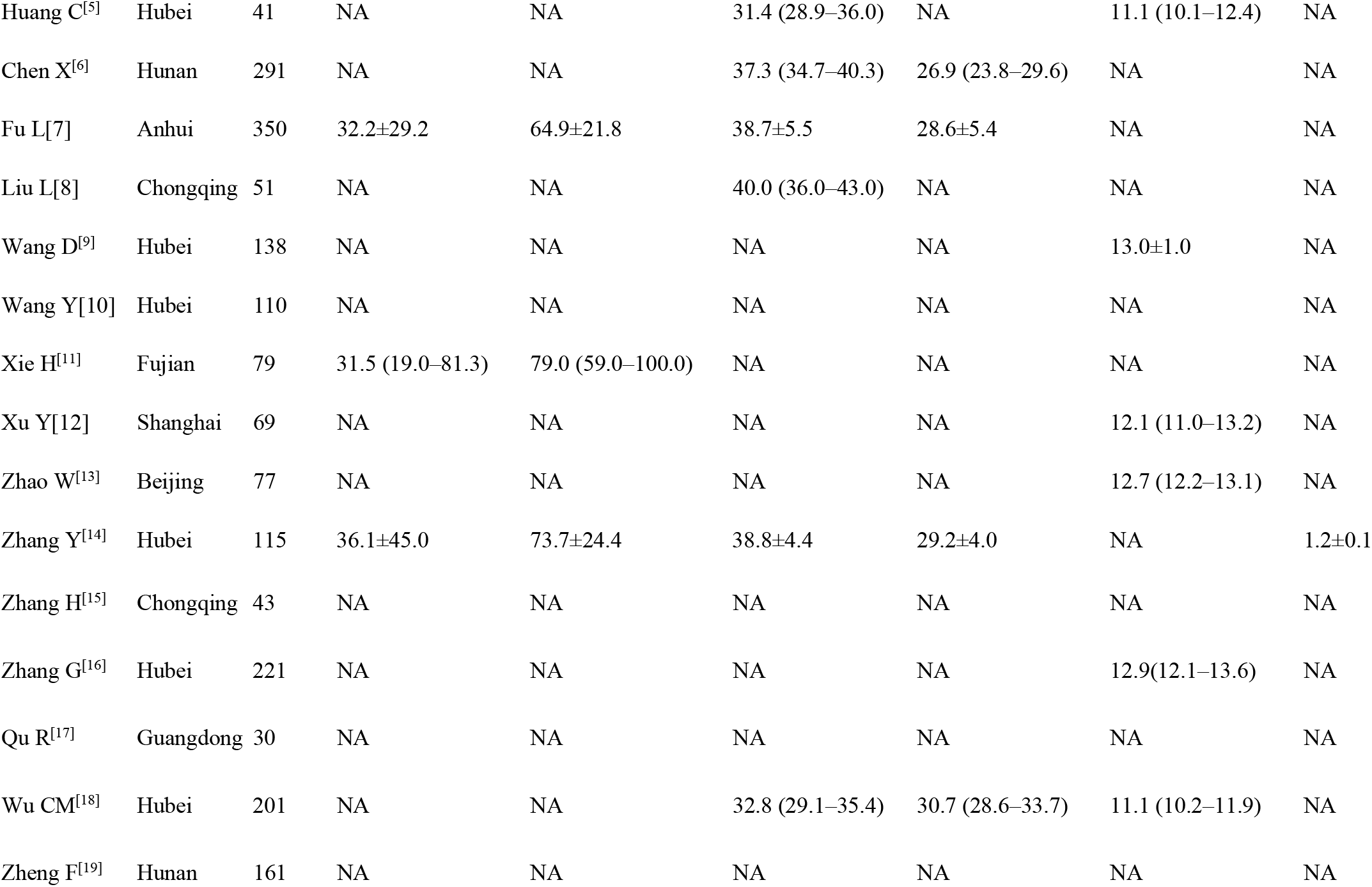
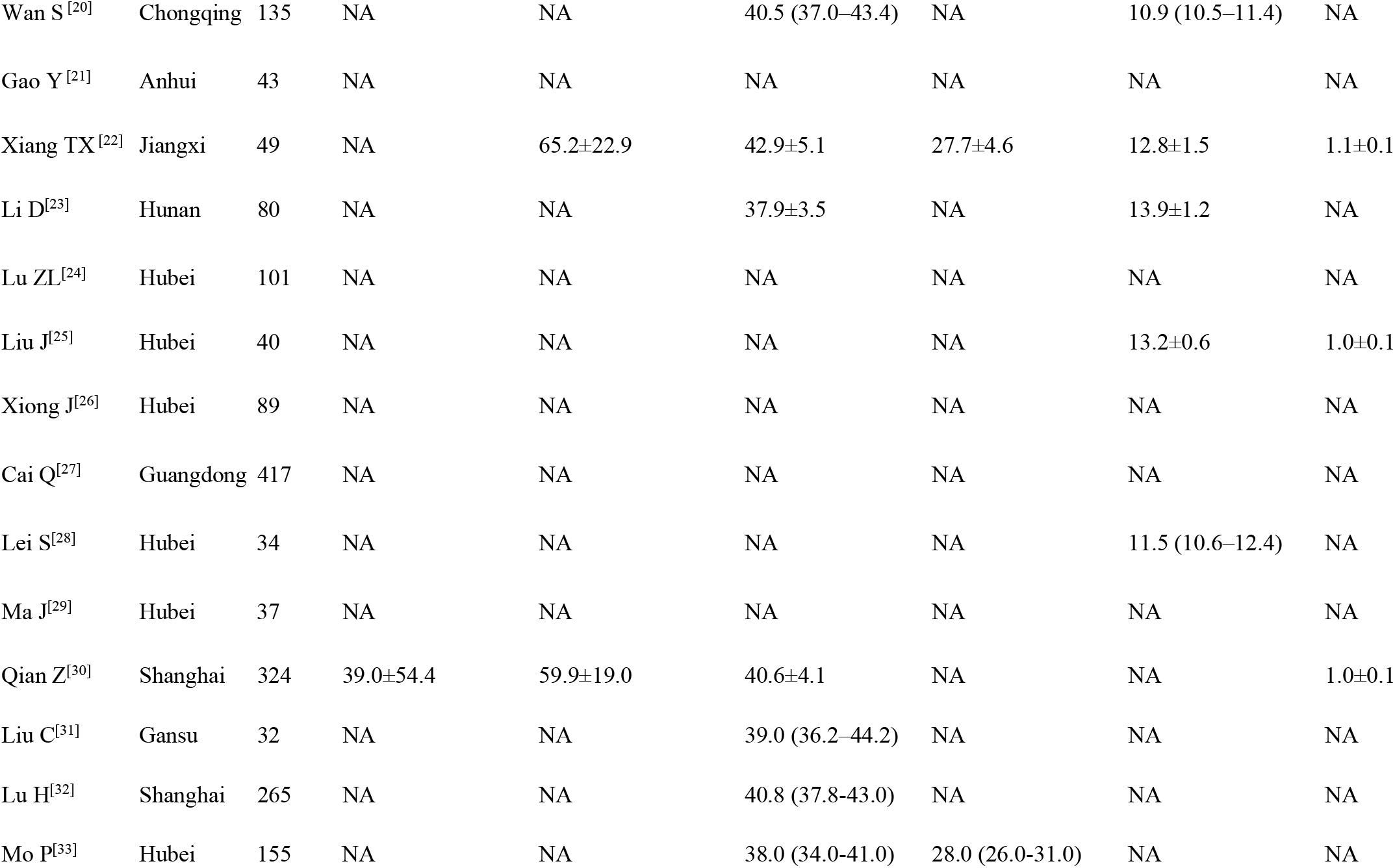
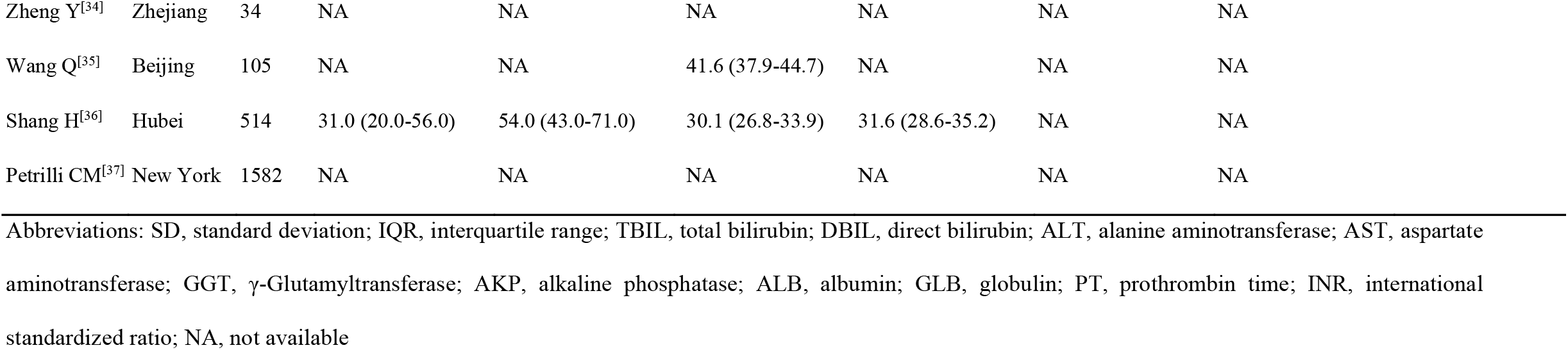
Characteristics of the enrolled studies for meta-analysis

**Figure 1.**
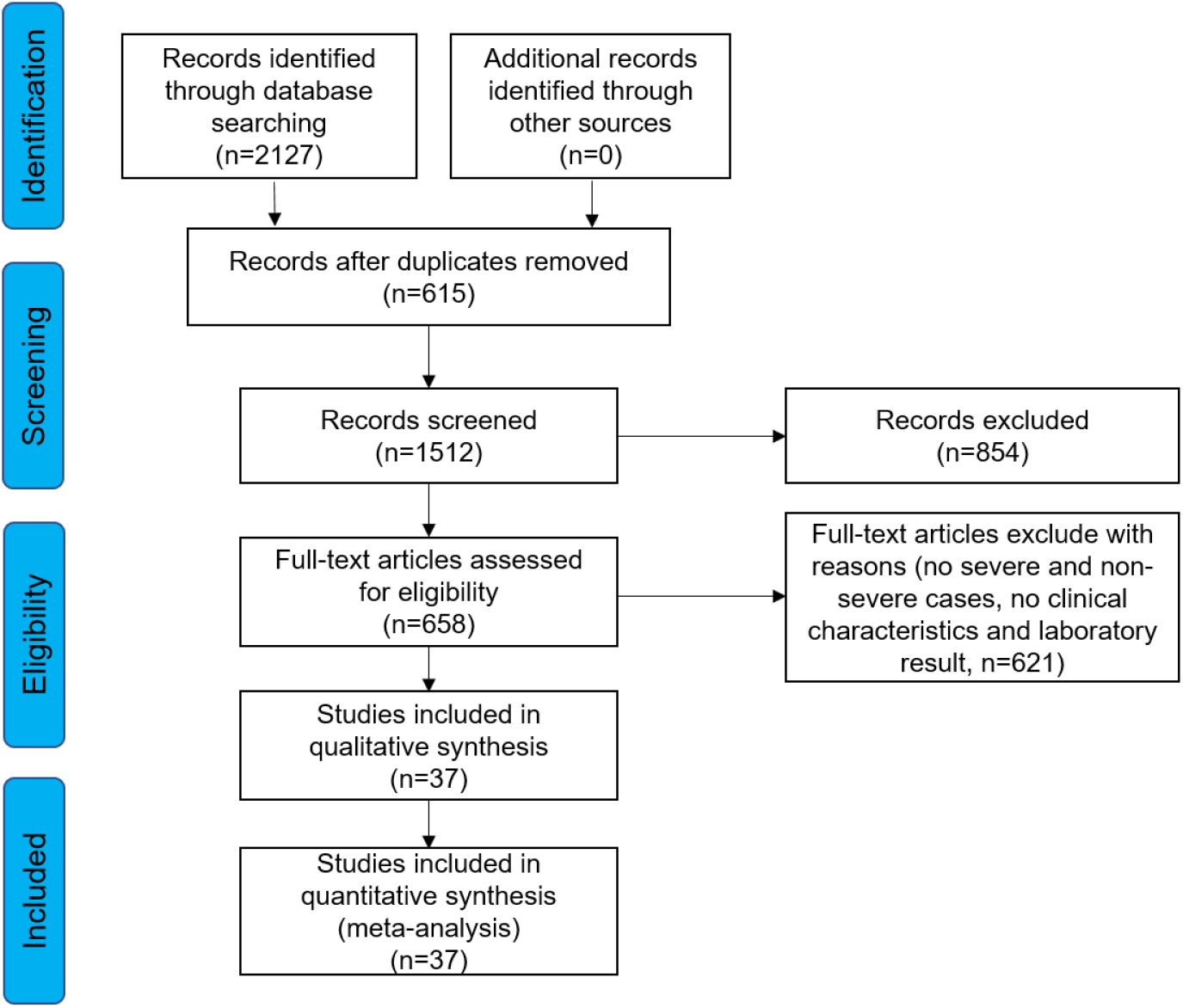
Study selection flow diagram. If all the liver chemistry indexes were not reported, these were regarded as “not available” and excluded from the meta-analysis. However, this study still enrolled studies that reported individual liver chemistry indexes if the severity of COVID-19 was reported in relation with the index.

### Hepatocellular injury-related abnormalities in liver chemistries

Of the enrolled studies, 37 reported assays of ALT or AST in a total of 6,235 patients with COVID-19. All but one of these studies were from China. Of the 36 enrolled studies from China, 16 (44.4%) were from Hubei Province, where Wuhan is located. The pooled mean ALT level was 36.4 IU/L in the patients with severe COVID-19 and 27.8 IU/L in the patients with non-severe COVID-19 (95% CI: − 9.4 to − 5.1, p < 0.0001; Figure 2A), with significant heterogeneity among the studies (*I*^2^ = 69%, p < 0.01). Similarly, the pooled mean AST level was 46.8 IU/L in the severe cases and 30.4 IU/L in the non-severe cases (95% CI: − 15.1 to − 10.4, p < 0.0001; Figure 2B). Significant heterogeneity was observed for the AST levels among the studies (*I*^2^ = 68%, p < 0.01), which was slightly lower than that of the ALT levels (*I*^2^ = 69%, p < 0.01). Potential publication bias was evaluated using a funnel plot (Supplementary Figure 2). In the patients with COVID-19, the mean AST level tended to be higher than the mean ALT level in both the severe and non-severe groups. Moreover, the gap between the AST and ALT levels (46.8 and 36.5 IU/L, respectively) was even more significant in the severe group (Figure 3). The evaluation of the publication bias is presented in Supplementary Figure 3.

**Figure 2.**
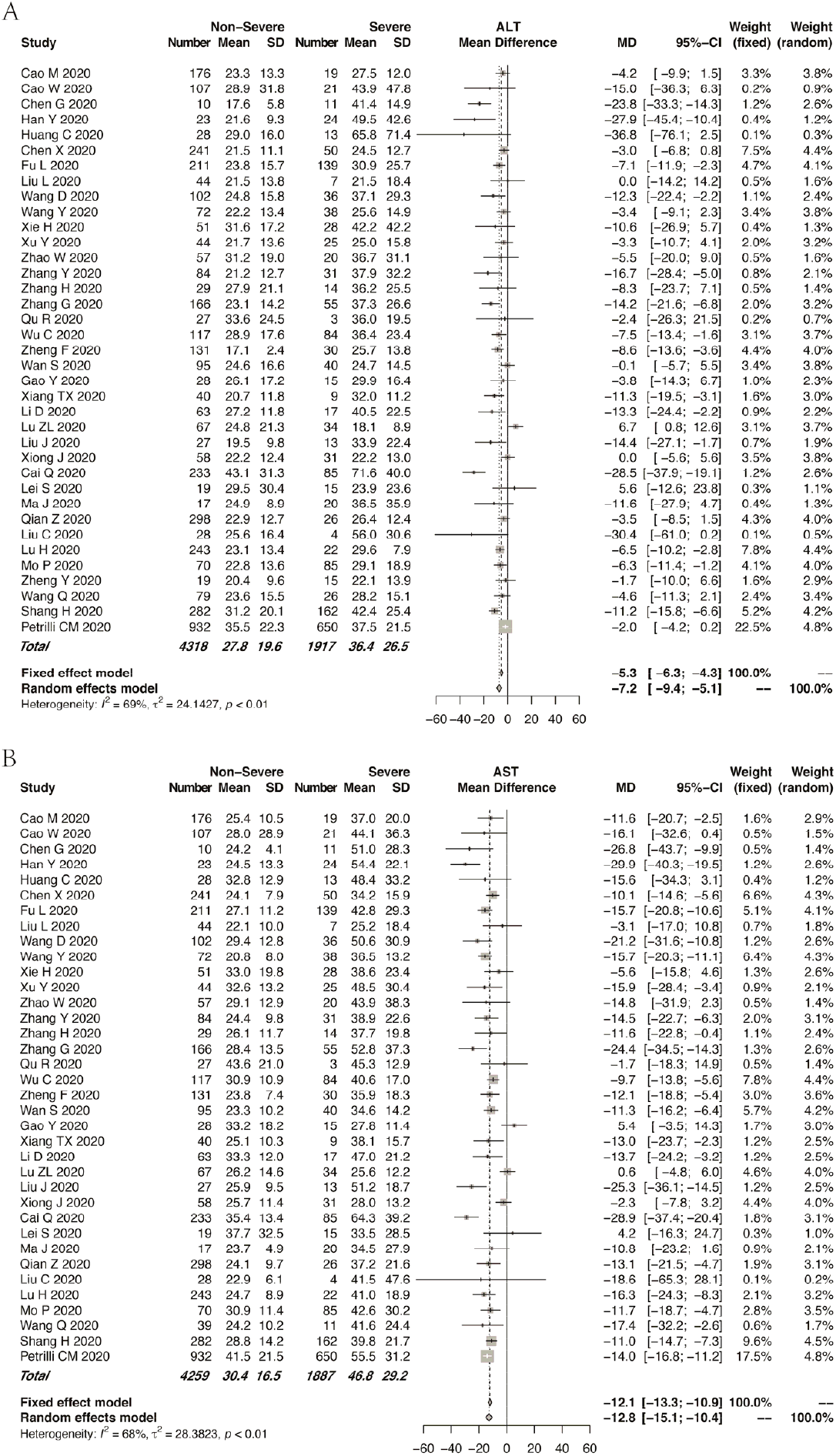
Forest plot of the association between serum ALT/AST level and disease severity. Pooled levels of (A) ALT and (B) AST in the patients with COVID-19. Abbreviations: ALT, alanine aminotransferase; AST, aspartate aminotransferase.

**Figure 3.**
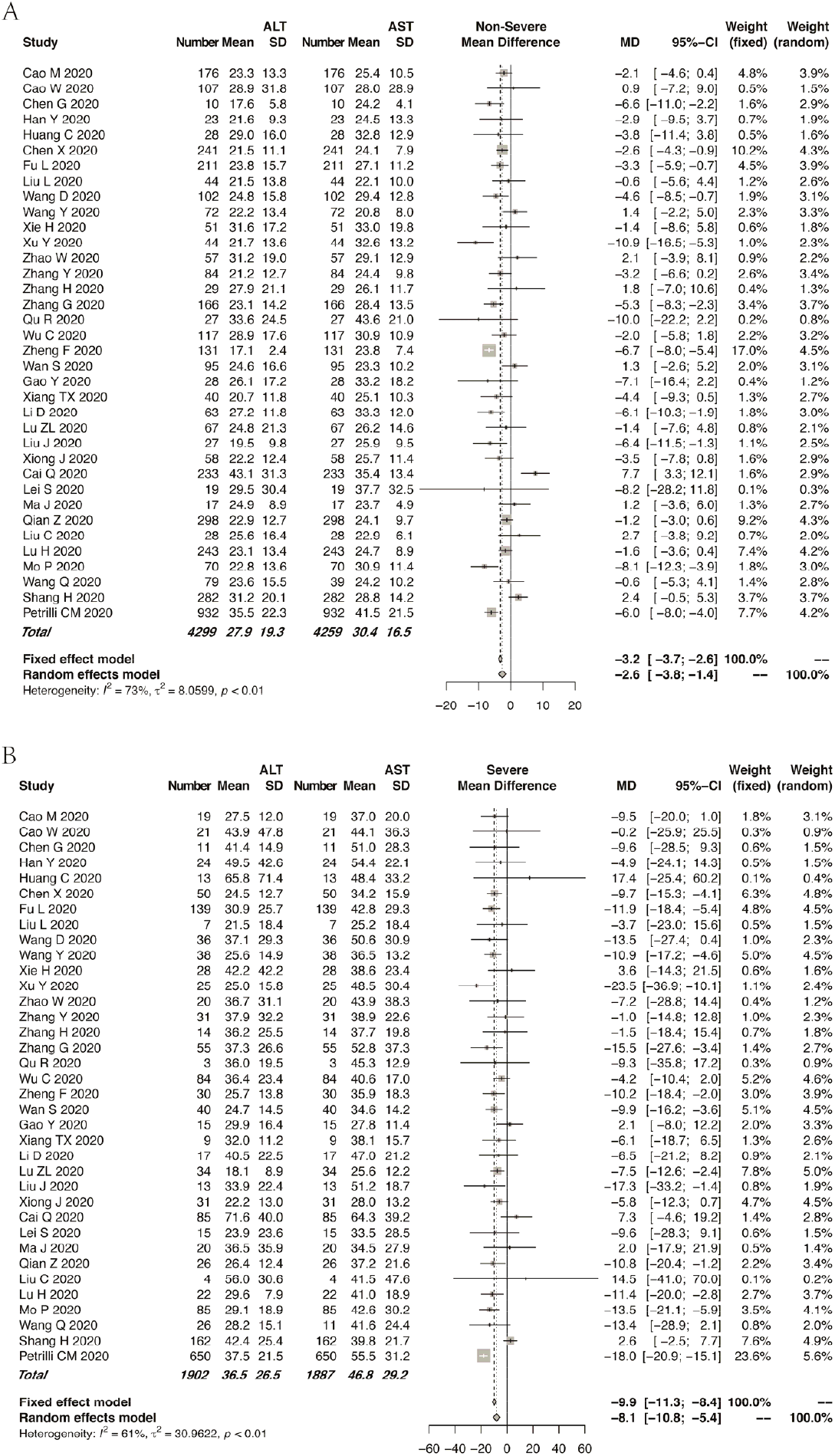
Forest plot for the comparison of ALT and AST levels in the patients with COVID-19 stratified by disease severity. A, Forest plot for the comparison of ALT and AST levels in the non-severe cases of COVID-19. B, Forest plot for the comparison of ALT and AST levels in the severe cases of COVID-19. Abbreviations: ALT, alanine aminotransferase; AST, aspartate aminotransferase.

### Cholestasis-related abnormalities in liver chemistries

Compared with the studies that reported ALT and AST levels, rather fewer studies focused on cholestasis-related indexes such as AKP, GGT, and DBIL levels. Of the enrolled studies for the meta-analysis, 8 reported AKP assays and 5 studies reported GGT measurements. The pooled mean AKP level was 66.2 IU/L in the patients with severe COVID-19 and 61.0 IU/L in those with non-severe COVID-19 (95% CI: − 11.2 to 0.1, p > 0.05; Figure 4A). The pooled mean GGT level was 43.2 IU/L in the severe group and 30.7 IU/L in the non-severe group (Figure 4B). The pooled mean TBIL level in the severe group was slightly higher than that in the non-severe group. However, the mean TBIL levels in both groups remained within the normal range (Figure 4C). Even fewer studies reported DBIL values in patients with COVID-19. In fact, no significant difference in mean DBIL level was found between the 2 groups (Figure 4D). For the TBIL levels, low heterogeneity was observed among the studies (*I*^2^ = 14%, p = 0.26). A funnel plot for the TBIL levels is shown in Supplementary Figure 4.

**Figure 4.**
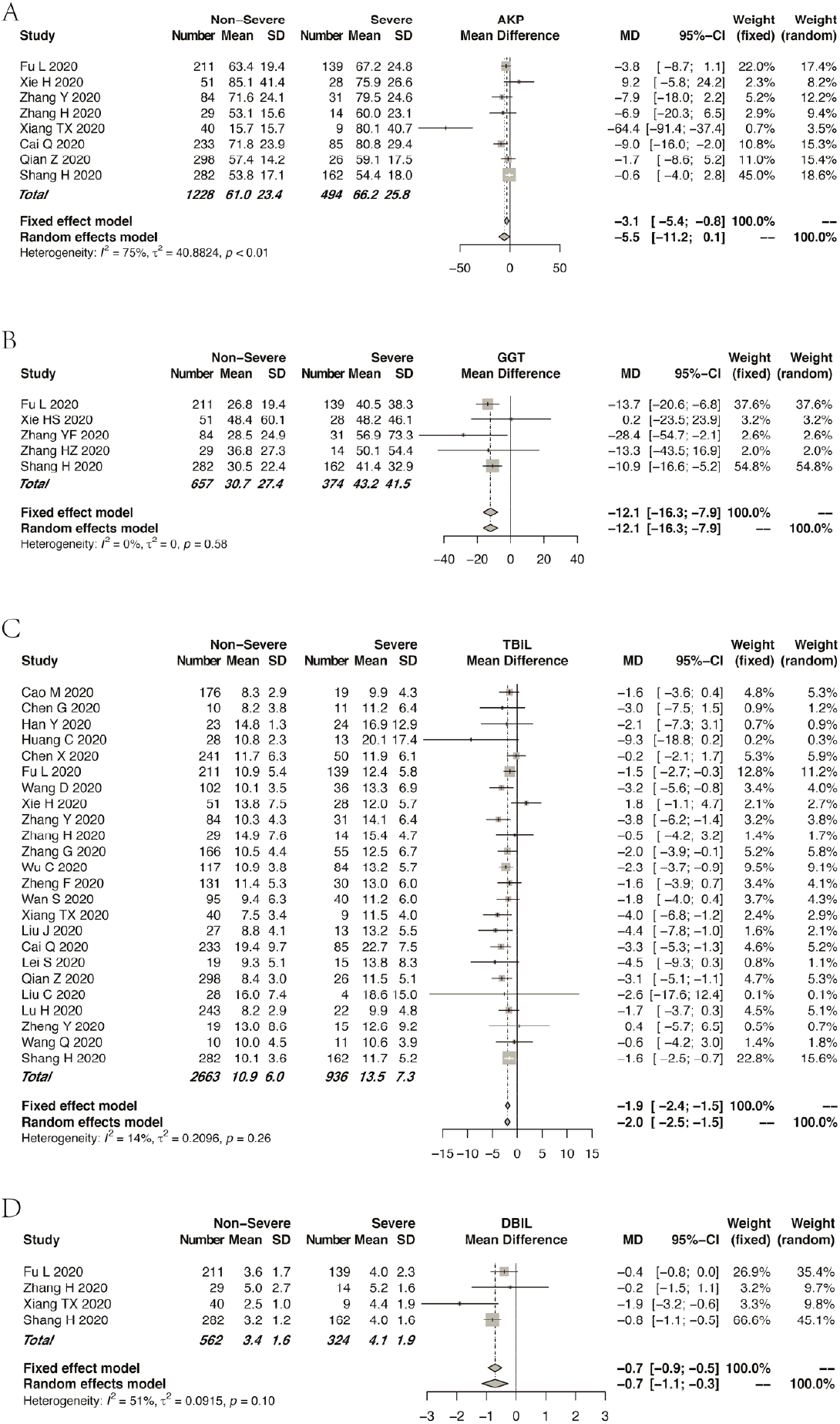
Forest plot for the association of the cholestasis-related indexes and disease severity. Pooled levels of (A) AKP, (B) GGT, (C) TBIL, and (D) DBIL in the patients with COVID-19. Abbreviations: AKP, alkaline phosphatase; GGT, γ-Glutamyltransferase; TBIL, total bilirubin; DBIL, direct bilirubin.

### Hepatocellular function-related abnormalities in liver chemistries

Sixteen studies compared the mean ALB levels according to COVID-19 severity, between 713 and 1,772 severe and non-severe cases, respectively (Figure 5A). A significant heterogeneity was observed among the studies (*I*^2^ = 61%, p < 0.01). The mean ALB level in the patients with severe disease was significantly lower than that in the patients with non-severe disease. No significant difference in GLB level was found between the groups (p = 0.17; Figure 5B). However, in the coagulation-related indexes such as PT and INR, no significant differences were found between the severity groups. The patients in the severe group tended to have longer PT or higher INR (Figure 5C, 5D). An evaluation of publication bias in relation to ALB level and PT is shown in Supplementary Figure 5.

**Figure 5.**
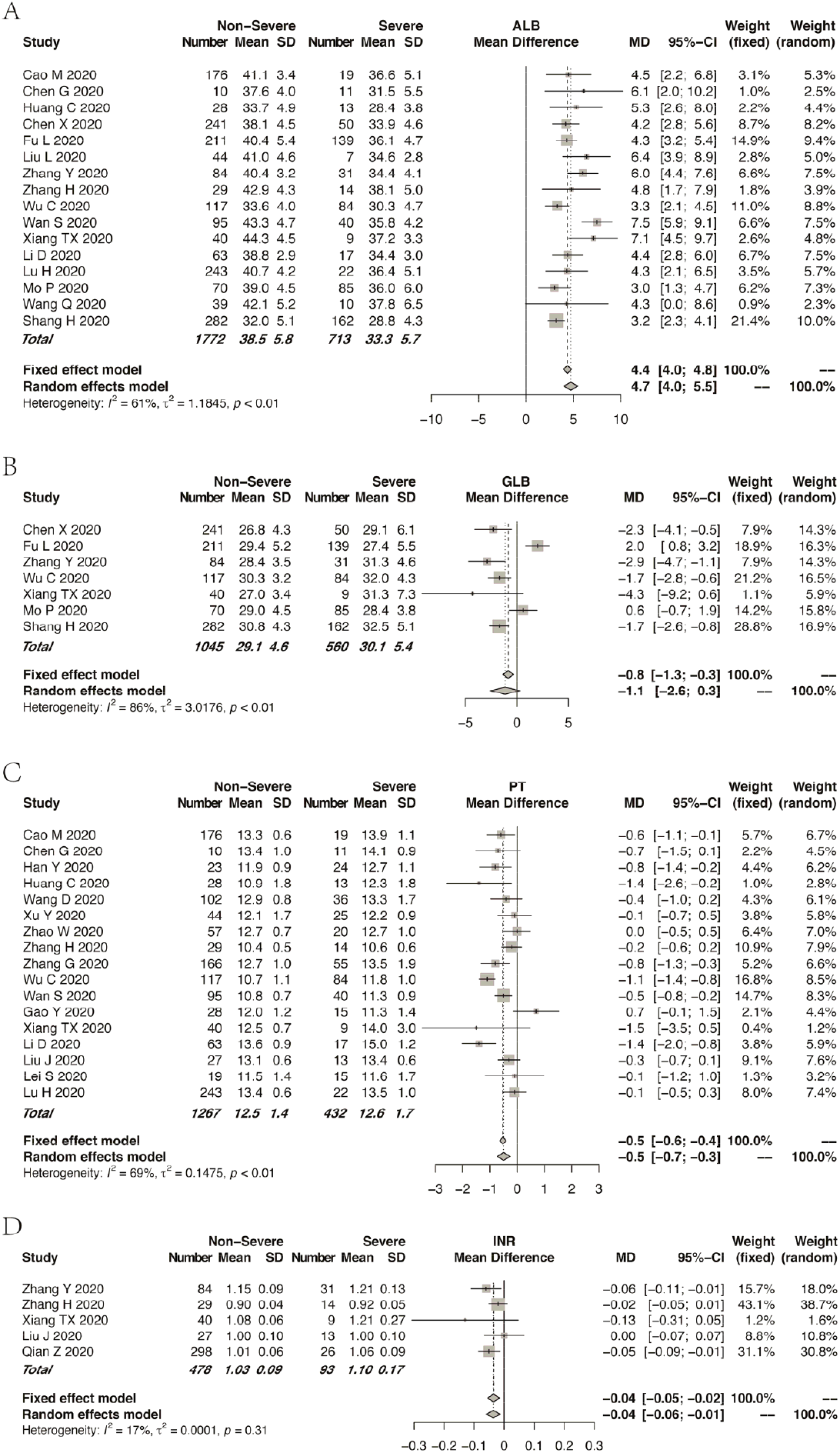
Forest plot for the association of the synthetic function-related indexes and disease severity. Pooled (A) ALB levels, (B) GLB levels, (C) PTs, and (D) INRs in the patients with COVID-19. Abbreviations: ALB, albumin; GLB, globulin; PT, prothrombin time; INR, international standardized ratio.

### DISCUSSION

In this meta-analysis, 37 studies that consisted a total of 6,235 patients with COVID-19 from China and the United States were enrolled. According to the pooled analysis, three patterns of liver impairment, namely hepatocellular injury, cholestasis, and hepatocellular dysfunction, can develop in a proportion of patients with COVID-19 at variable severity. In brief, the patients with severe COVID-19 tended to have higher ALT/AST, AKP/GGT, and TBIL levels; higher INR; and prolonged PT. However, the severe cases had lower ALB levels than the non-severe cases. The patients with COVID-19 often had higher AST levels than ALT levels, especially in severe cases. We also observed a tendency of the severe cases to arise in the elderly.

Although the liver may act as the latent target of SARS-CoV-2, the actual prevalence of abnormal liver chemistries could be underestimated because many studies did not report cholestasis-related indexes such as AKP and GGT levels, and synthetic function-related indexes such as ALB level and INR. Moreover, most studies reported ALT/AST levels on the day of admission but not throughout the disease course. This issue further compromises the role of liver chemistries in disease monitoring and provides an early warning against severe cases. As SARS-CoV-2 can lead to bile duct damage by conquering the ACE2 expressed on cholangiocytes and induce a subsequent cholestatic liver injury [8], cholestasis-related abnormalities could be overlooked.

Cumulating studies have linked abnormal liver chemistries to the severity of COVID-19 [9, 19, 25]. Patients with abnormal liver test results are at higher risk of progression to severe disease [9]. In fact, coronavirus infection can cause direct damage to liver cells [11]. Moreover, several underlying diseases, comorbidities, and complications that develop in the course of the disease, such as sepsis and multiple-organ failure, and drugs that can cause potential liver damage also increase the risk of liver injury. Lopinavir/ritonavir use during hospitalization has been reported to possible lead to liver damage [9, 52]. The liver chemistry tests in the enrolled studies were all performed on admission, which suggests that the influences of the drugs on the liver tests, if any, should be minor.

ALB level and PT are known to reflect hepatocellular function. Albumin, which has a circulating half-life of 3 weeks, is a plasma protein exclusively synthesized by the liver [53]. Hypoalbuminemia results from and reflects the inflammatory state, which leads to inflammatory exudate. Effective nutrition support helps to correct hypoalbuminemia [54]. Our meta-analysis revealed that ALB level was lower in the severe cases than in the non-severe cases, which indicated that the severe cases tended to have more intense inflammation and require more solid nutrition support. PT is a far more sensitive measure of hepatocellular function than ALB level because PT may be prolonged in patients with severe liver disease duration of <24 h [14]. In accordance with the alteration of the ALB level, PT was prolonged in the severe cases, which further indicated impairment of hepatocellular function in the severe cases [53]. According to our meta-analysis, another interesting feature of liver impairment related to COVID-19 is that the AST level often overrides the ALT level, especially in severe cases. By contrast, in patients with chronic hepatitis B or nonalcoholic fatty liver disease, the ALT level is generally higher than the AST level. Although ALT is present primarily in the liver and is a more specific marker of hepatocellular injury, the distribution of AST is far wider than that of ALT, including the cardiac muscle, skeletal muscle, kidney, and brain [14]. An elevated AST level accompanied by a normal ALT level often suggests cardiac or muscle disease. In fact, cardiac injury is frequent in severe cases of COVID-19, especially in deceased patients [25, 55].

This study has some substantial merits. First, this meta-analysis comprehensively summarized the rapidly evolving and sometimes confusing literature on COVID-19 regarding the manifestation of liver chemistries. The extensive coverage of 37 studies allowed a more precise evaluation of the abnormalities of liver chemistries. Our subgroup analysis revealed that the abnormal liver chemistries were associated with a more severe disease course, highlighting the importance of a more extensive monitoring of liver chemistries for both diagnostic and prognostic purposes. Second, this analysis extensively covered all three patterns of liver impairment, namely hepatocellular injury, cholestasis, and hepatocellular dysfunction. Most observations focused on ALT, AST, and ALB levels. However, cholestasis-related impairment (e.g., abnormal AKP and GGT levels) tended to be inadvertently ignored. Moreover, we also compared hepatocellular dysfunction between the severe and non-severe cases. The alarmingly high prevalence of hypoalbuminemia in the severe cases prompts further nutrition support in severe cases. In addition, coagulation dysfunction in severe cases requires vigilance. Third, the enrolled studies included multiple observations not only from mainland China but also from other ethnic groups. This facilitates the assessment of abnormal liver chemistries related to COVID-19 in a broader ethnic context. Fourth, eligible studies preprinted in medRxiv and bioRxiv were also covered, which ensured that our analysis has a clear leading position. However, our study has a few limitations. As mentioned earlier, cholestasis-related indexes such as AKP/GGT level may be under-reported in quite a number of studies, which may lead to less precise pooled data. Second, most studies came from mainland China, which precluded a more precise estimate of the abnormalities of liver chemistries in the other ethnic groups. Most of the studies that came from mainland China seem to have an adverse impact. On the contrary, this helps to abate the heterogeneity caused by the disease grouping, as some potential discrepancies may exist in the definition of severe and non-severe cases of COVID-19 between different countries.

## CONCLUSION

In this meta-analysis, we comprehensively described three patterns of liver impairment related to COVID-19, namely hepatocellular injury, cholestasis, and hepatocellular dysfunction, according to COVID-19 severity. Patients with abnormal liver test results are at higher risk of progression to severe disease. Close monitoring of liver chemistries provides an early warning against disease progression.

## Data Availability

NA

## Guarantor of the article

Professor Jin-Shui Pan

## Specific author contributions

JSP and MZH were involved with the study conceptualization and design; analysis and interpretation of data; drafting of the manuscript; and approval of the final version of the manuscript; XD, DYZ, YYC, WMC, QQX, and YDR were involved in data retrieval.

## SUPPLEMENTARY FIGURE LEGENDS

Supplementary Figure 1. Association between age and disease severity

A, Forest plot for the association of age and disease severity. B, Funnel plot for the association of age and disease severity.

Supplementary Figure 2. Funnel plot for the association of ALT/AST level and disease severity

A, Funnel plot for the association of ALT level and disease severity. B, Funnel plot for the association of AST level and disease severity.

Abbreviations: ALT, alanine aminotransferase; AST, aspartate aminotransferase.

Supplementary Figure 3. Funnel plot for the comparison between ALT and AST levels stratified by disease severity

A, Funnel plot for the comparison of ALT and AST levels in the non-severe cases. B, Funnel plot for the comparison between ALT and AST level in the severe cases.

Abbreviations: ALT, alanine aminotransferase; AST, aspartate aminotransferase.

Supplementary Figure 4. Funnel plot for the association of TBIL level and disease severity

Abbreviations: TBIL, total bilirubin.

Supplementary Figure 5. Funnel plot for the association of ALB level/PT and disease severity

A, Funnel plot for the association of ALB level and disease severity. B, Funnel plot for the association of PT and disease severity. Abbreviations: ALB, albumin; PT, prothrombin time.

